# Retrospective and prospective studies evaluating the performance of the SARS-Cov-2 “AQ+ COVID-19 Ag Rapid Test” from InTec on symptomatic and non-symptomatic patients

**DOI:** 10.1101/2022.09.29.22280448

**Authors:** Thierry Prazuck, Raphael Serreau, Aurelie Theillay, Sandra Pallay, Daniela Pires-Roteia, Fanny Prazuck, Fabien Lesne

## Abstract

For the last two years, the SARS-CoV-2 virus spread all around the world and led to the COVID-19 pandemic. The need of methods to control the pandemic and to propose rapid and efficient diagnostic tools has emerged. In this perspective, SARS-CoV-2 rapid antigen detection tests (RADT) have been developed. We performed a retrospective study on 638 collected nasopharyngeal samples used for reference RT-qPCR diagnosis to compare the AQ+ COVID-19 Ag Rapid Test” from InTec (AQ+ InTec test) performance with other commercially available RADT. We analysed the sensitivity and specificity of the different tests and showed a better overall performance of the AQ+ InTec test, which was confirmed on the SARS-Cov-2 Omicron variant. We then conducted a prospective study on 1428 patients, to evaluate the sensitivity and specificity of the AQ+ InTec test on nasal and nasopharyngeal samples in a point of care setting. We showed that sensitivity and specificity reach acceptable criteria regarding the official recommendations of the MDCG 2021-21 in both symptomatic and asymptomatic patients. Overall, the results of these two studies confirm that the AQ+ InTec test is a valuable tool for testing in a pandemic context with a high proportion of asymptomatic patients who are potential carriers for the SARS-CoV-2 virus and is performant on the most current circulating variant Omicron.

**Highlights:**

- The sensitivity of the AQ+ COVID-19 Ag Rapid Test from InTec reaches 94.4% on nasal samples and 97.4% on nasopharyngeal samples.
- The performance of the test remains high on asymptomatic patients with a sensitivity of 89.2% on nasal samples and 97.0% on nasopharyngeal samples.
- Prospective study in a point of care setting revealed a better sensitivity compared with other commercially available rapid antigen detection tests

## Introduction

In late 2019, in Wuhan China, a new type of respiratory infection caused by a novel coronavirus strain, the SARS-CoV-2, emerged. The disease was named COVID-19 for CoronaVirus Disease 2019. Since then, it has spread to the entire global population and was given the pandemic status on March 11, 2020 by the World Health Organization (WHO)^1^.

SARS-CoV-2 infection mainly causes pneumonia and upper/lower respiratory tract infection^2^. Symptoms of COVID-19 infection appear after an average incubation period of 5.2 days^2^. The most common early symptoms of COVID-19 disease are fever, cough, and fatigue, but other symptoms include headache, sore throat, and even acute respiratory distress syndrome, leading to respiratory failure.

The pandemic has become a major public health problem worldwide due to the large number of COVID-19 patients requiring hospitalization and breathing assistance. This exceptional influx of patients during peak epidemics exceeds the capacity of national health systems leading to global hospital overloads. Therefore, a rapid and accurate identification of patients requiring supportive care and isolation is important for the management of COVID-19.

Along with a massive vaccination program, highly sensitive and specific tests are crucial to identify and manage COVID-19 patients and implement control measures to limit the pandemic. The current gold standard test is a molecular identification of the genetic material of the virus by Reverse Transcription-Polymerase Chain Reaction (RT-qPCR) from a nasopharyngeal swab sample^3,4^. This test is very sensitive and reliable, but its generalisation is hampered by the fact that samples can only be taken by a professional trained in the procedure and requires a laboratory and expensive equipment.

New rapid virus antigen detection tests (RADT) are part of the “Test-Alert-Protect” strategy^5^. These tests are rapid, inexpensive, easy to use, with an estimated reading time of 15 minutes. In 2020, the French National Authority for Health (HAS) published a favourable opinion on RADT for the diagnosis of symptomatic patients up to 7 days after the onset of symptoms, as an alternative to RT-qPCR on nasopharyngeal swabs, and for asymptomatic individuals in the context of large-scale operations, targeting high-risk of infection populations ^6,7^. Furthermore, the HAS supports the deployment of antigenic self-tests based on oropharyngeal or nasal swabs as a complementary tool in the SARS-CoV-2 screening strategy^8^. InTec PRODUCTS has developed the RADT AQ+ COVID-19 Ag Rapid Test (AQ+ InTec test). Two studies have recently been conducted to evaluate the test performance using the nasal and nasopharyngeal sampling method and the European Guidelines for validation of COVID-19 RADTs^9,10^.

First, a retrospective study aimed at comparing the sensitivity of the AQ+ InTec test with other commercial SARS-CoV-2 RADT, using collected nasopharyngeal samples for reference testing with RT-qPCR. Then, a prospective study evaluated the AQ+ InTec test in a point of care setting. To this aim, the sensitivity of the test in both nasal and nasopharyngeal samples was analysed and a comparison with the results of the reference RT-qPCR test depending on the CT level was made. Moreover, the sensitivity of the test was evaluated regarding the symptom status of the patients. Finally, the specificity of the test was also evaluated.

## Material and methods

### Retrospective study

The study was approved by the Orleans regional Hospital ethics committee and notified to the French data protection authority. The study was retrospective, on collected samples and aiming at validating the performances (sensitivity and specificity) of the “AQ+ COVID-19 Ag Rapid Test” (InTec), compared to the reference method (RT-qPCR) and comparative antigen detection-based tests.

#### Reference method

The reference method was a RT-qPCR test performed on nasopharyngeal sample in subjects who came to the investigative center to perform a COVID-19 molecular test (RT-qPCR, nasopharyngeal swab) with or without symptoms.

The RT-qPCR test for SARS-CoV-2 was performed in the virology unit of the CHR Orléans, France. Nucleic acid extraction was performed with an automated sample preparation system MGISP-960 (MGI, China). Real-time PCR detection of SARS-CoV-2 RNA targeting the ORF1ab, S and N genes was performed with the TaqPath V2 Covid-19 Multiplex RT-PCR kit (Thermofisher). Amplification was performed on QuantStudio5 (Applied Biosystems). The results of the assay were analysed according to the manufacturer’s instructions. The assay includes an internal RNA extraction control and an amplification control. The samples were analysed considering the new positivity criteria of the French Microbiology Society’s expert committee ^11^, in particular considering the specific characteristics of the Thermofisher kit used for the RT-qPCR measurement.

#### AQ+ COVID-19 Ag Rapid Test (InTec) and Comparative rapid tests

The AQ+ COVID-19 Ag Rapid Test (GJ22020185 (25T/Kit)) is a colloidal gold immunochromatographic test for the qualitative detection of SARS-CoV-2 core antigens potentially present in human nasal swabs from individuals suspected of having COVID-19 within the first 7 days of symptom onset or that without symptoms. The test is used as an aid in the diagnosis of SARS-CoV-2 infection. It is suitable for use under healthcare professional supervision. Remote healthcare professional supervision can be used with appropriate clinical governance once training has been completed and verified.

If SARS-CoV-2 antigens are present in the sample, an antibody-antigen complex will form upon contact with anti-SARS-CoV-2 monoclonal antibodies conjugated with coloured particles. This coloured complex migrates by capillary action across the membrane to the test line (T). If no SARS-CoV-2 antigen is present in the sample, no colour appears on the test line (T). The control line (C) is a control that should appear if the test procedures are performed correctly.

The following RADT were used as comparatives: Abbott Panbio, Roche SARS-CoV-2 Rapid Antigen Test (SD Biosensor) and Siemens Clinitest (Orientgene).

After performing the RT-qPCR reference test, nasopharyngeal leftover samples were frozen in Viral Transport Media (VTM), (Dasky), then used to perform the AQ+ COVID-19 Ag Rapid Test and the comparatives, according to each manufacturer recommendations.

#### Limit Of Detection (LOD) analysis

LOD is defined as the lowest concentration of the analyte that can be reliably detected. Serial dilutions were made of the sample collected in VTM, using the leftovers after RT-qPCR reference testing (from 1,00E+06 to 3,20E+01 gcn/mL). Independent swabs were immersed in the diluted sample collected in the VTM, and the RADT test previously described were performed according to each manufacturer instructions (InTec, Orient Gene, Roche, Abbott).

### Prospective study

The study was approved in April 2022 by the Sud Ouest et Outre Mer ethics committee (N° 2022-A00503040) and was notified to French data protection authority. In accordance with the Declaration of Helsinki, all adult participants or legal representative for participating children provided written informed consent before undergoing any study-specific procedure. The study was prospective, aiming at validating the performances (sensitivity and specificity) of the “ AQ+ COVID-19 Ag Rapid Test “ (InTec), compared to the reference method (RT-qPCR).

#### Reference method

The reference method was a RT-qPCR test performed, as previously described, on nasopharyngeal sample in subject obtained during the participant’s screening visit (as part of standard of care)

#### AQ+ COVID-19 Ag Rapid Test (InTec)

The A AQ+ COVID-19 Ag Rapid Test Test (InTec) was performed on fresh nasal or nasopharyngeal swab collected in subject obtained during the participant’s visit for RT-qPCR testing, according to manufacturer instructions.

## Results

### Retrospective study

Sample collection was performed from subjects who came to the investigative center to perform a COVID-19 molecular test (RT-qPCR, nasopharyngeal swab) with or without symptoms. After testing, sample leftovers were frozen and subsequently used to carry out the AQ+ InTec test and the commercial comparatives RADT. The study was conducted on 638 samples tested with the reference RT-qPCR method: 329 were negative and 309 were positive.

The results obtained with the AQ+ InTec test were compared with other SARS-CoV-2 RADT from Orientgene, Roche and Abott.

The sensitivity of the AQ+ InTec test was higher, with the detection of 218 positive patients out of 309 (70.6%; compared to 168, 51 and 45 respectively with the Orient Gene, Roche, and Abbott tests). This difference of sensitivity was observed for both SARS-CoV-2 Omicron and Delta variants (Table 1, Figure 1). Of note, the difference between AQ+ InTec test and the other RADT used, especially Roche and Abbott’s, was more marked as the CT level was lower (Table S 1).

**Table 1.** Comparison analysis of sensibility and specificity of AQ+ InTec test compared with other brand antigenic based rapid tests

| Rapid tests' brands |  | Intec (1) | Orient Gene (2) | Roche (3) | Abbott (4) |
| --- | --- | --- | --- | --- | --- |
| Sensitivity | Overall (n=309) | 70.6%<br>[65.5-75.6] | 51.1%<br>[45.6-56.7] | 19.7%<br>[15.3-24.2] | 14.6%<br>[10.7-18.5] |
|  | Omicron (n=249) | 73.9%<br>[68.4-79.3] | 53.4%<br>[47.2-59.7] | 20.9%<br>[15.8-25.9] | 15.3%<br>[10.8-19.7] |
|  | Delta (n=51) | 51%<br>[37.3-64.7] | 33.3 %<br>[20.4-46.3] | 15.7%<br>[5.7-25.7] | 11.8%<br>[2.9-20.6] |
| Specificity | Overall (n=309) | 100% | 100% | 100% | 100% |

Statistical test on overall sensitivity: P value 1 vs 2: <0.0001; P value 2 vs 3: <0.0001; P value 3 vs 4: non-significant Chi2: 274; Global p value: P<0.0001

**Figure 1.**
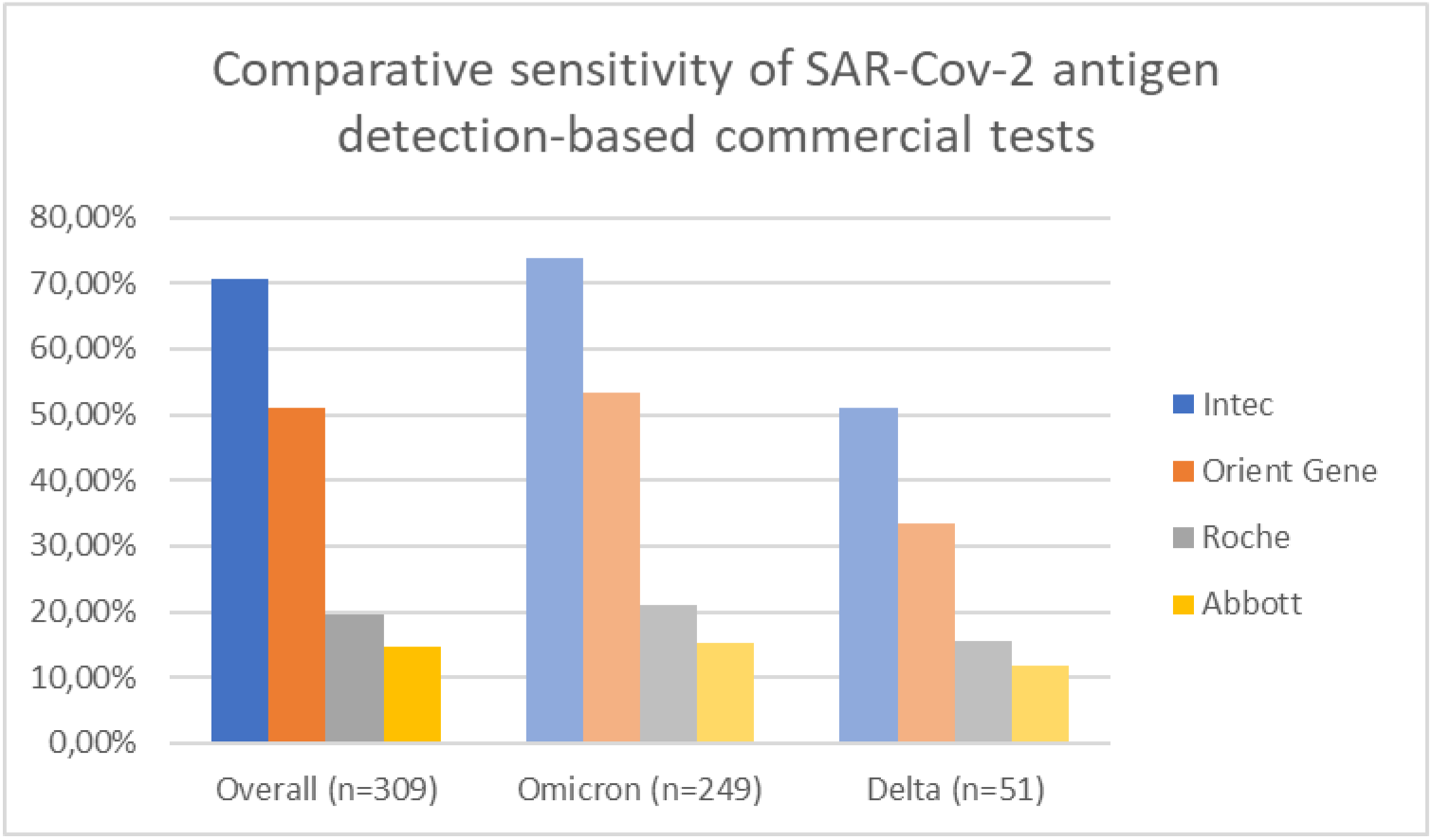
Comparative sensitivity of SARS-CoV-2 antigen detection-based commercial tests

**Figure 2:**
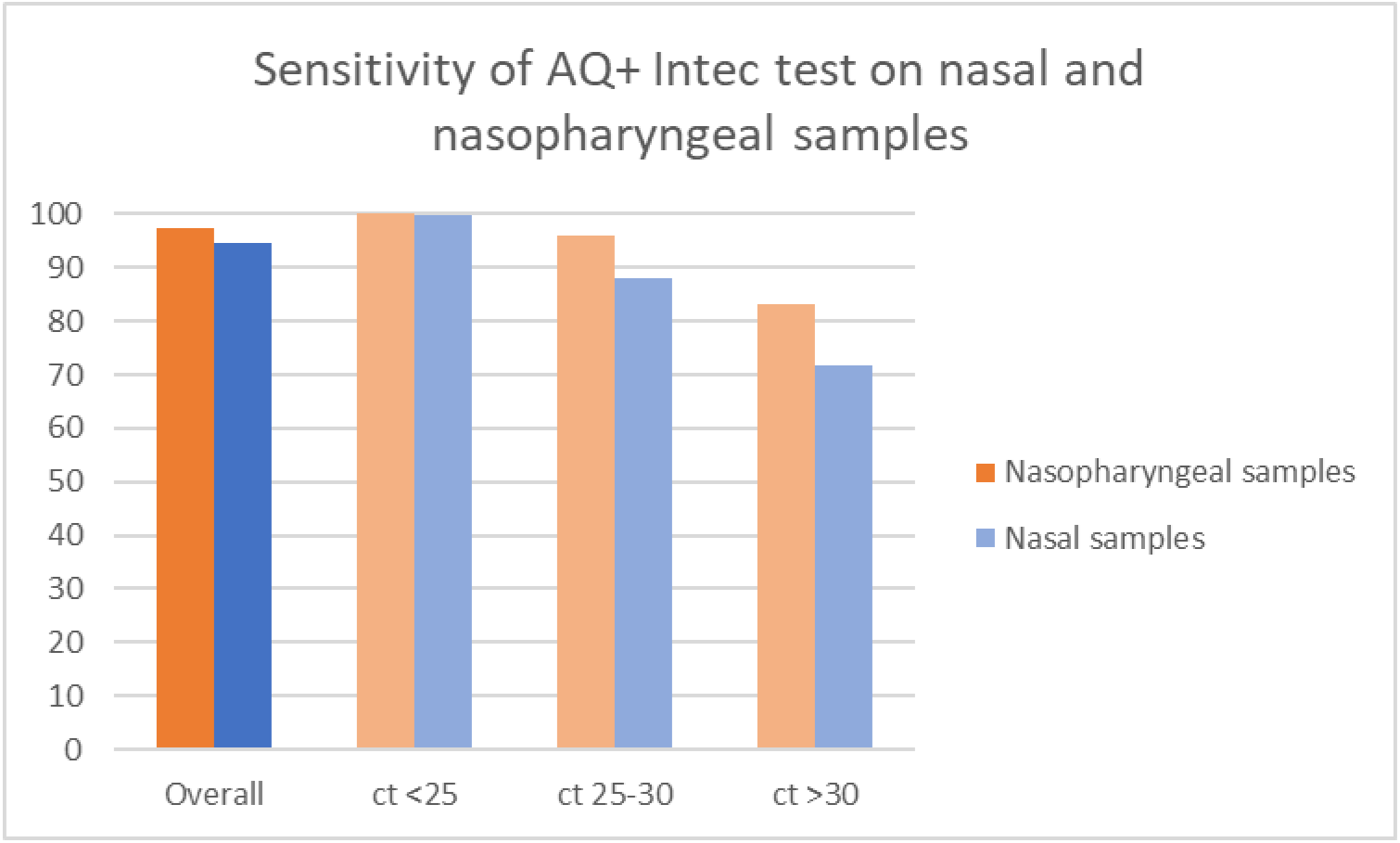
Sensitivity of the AQ+ InTec test on nasal and nasopharyngeal samples and corresponding CT levels of the reference RT-qPCR test.

Considering the conditions in which the sample was obtained (storage in VTM and freezing), the sensitivity of the AQ+ InTec test was quite high but did not reach the criteria defined in the MDCG 2021-21 ^12^ which places diagnostic sensitivity threshold higher than 80% (RADT) relative to the SARS-CoV-2 RT-qPCR test.

All the tests had a specificity of 100% (Table 1) and no difference was observed.

The limit of detection (LOD) of the AQ+ InTec test has been evaluated and compared with the three other RADT. The lower LOD was observed with the AQ+ InTec test (1.60E+02), followed by Orient Gene (2.00E+04).

In all, the AQ+ InTec test showed a higher sensitivity on samples from symptomatic and asymptomatic patients, compared to other RADT commercially available ^13–16^.

### Prospective study

Considering the promising results of the retrospective study, it was foreseen to evaluate the performance of the AQ+ InTec test, by trained professional in a point of care setting.

Overall, 660 samples were collected from patients and tested positive with the reference RT-qPCR. A nasal sample was taken from all patients and 378 of them underwent both a nasal and a nasopharyngeal sampling. The AQ+ InTec test sensitivity analysis was performed on 391 patients with known SARS-CoV-2 infection (nasal sample positive with RT-qPCR). The specificity analysis was performed on 258 patients with no SARS-CoV-2 infection (nasal sample negative with RT-qPCR).

#### AQ+ InTec test sensitivity: comparison with CT level of the reference test

To evaluate the performance of the AQ+ InTec test on nasal and nasopharyngeal samples, the results obtained with the test were compared to the reference RT-qPCR results. Two groups of patients were analysed: nasal and nasopharyngeal samples. In the first group, the sensitivity of the AQ+ InTec test was measured on nasal samples of 391 patients. Among these, SArS-Cov2 antigen was detected in 369 patients by the AQ+ InTec test compared with the reference method (RT-qPCR), with an overall sensitivity of 94.4% (95% CI [92.1-96.7]). Interestingly, the sensitivity was as high as 99.6% (95% CI [98.9 – 100.0]) when the CT level was below 25 in the reference test (Table 3, Figure 1).

**Table 2.** Limit of detection analysis

|  |  |  |  |  |  | Total<br>N=1 |
| --- | --- | --- | --- | --- | --- | --- |
|  |  | Intec | Orient Gene | Roche | Abbott | Estimated<br>gcn/mL |
| PCR CT (N<br>gene) | 25 | Positive | Positive | Negative | Negative | 1,00E+06 |
|  | 27 | Positive | Positive | Negative | Negative | 5,00E+05 |
|  | 29 | Positive | Positive | Negative | Negative | 1,00E+05 |
|  | 31 | Positive | Positive | Negative | Negative | 2,00E+04 |
|  | 32 | Positive | Negative | Negative | Negative | 4,00E+03 |
|  | 34 | Positive | Negative | Negative | Negative | 8,00E+02 |
|  | 36 | Positive | Negative | Negative | Negative | 1,60E+02 |
|  | Undet. | Negative | Negative | Negative | Negative | 3,20E+01 |

**Table 3:** Sensitivity of the AQ+ InTec test on nasal and nasopharyngeal samples and corresponding CT levels of the reference RT-qPCR test.

|  |  | Positive RT-qPCR<br>(nasopharyngeal) | Positive AQ+ Intec<br>test | Sensitivity<br>% [95% CI] |
| --- | --- | --- | --- | --- |
| Nasal samples |  |  |  |  |
| Overall |  | 391 | 369 | 94.4<br>[92.1-96.7] |
| CT level | <25 | 259 | 258 | 99.6<br>[98.9 – 100.0] |
|  | 25-30 | 100 | 88 | 88.0<br>[81.6 – 94.4] |
|  | >30 | 32 | 23 | 71.8<br>[56.3 – 87.5] |
| Nasopharyngeal samples |  |  |  |  |
| Overall |  | 269 | 262 | 97.4<br>[95.5-99.3] |
| CT level | <25 | 168 | 168 | 100<br>[100 – 100.0] |
|  | 25-30 | 77 | 74 | 96.1<br>[91.8– 100] |
|  | >30 | 24 | 20 | 83.3<br>[68.4– 98.2] |

The sensitivity of the AQ+ InTec test was also measured on nasopharyngeal samples of 269 patients detected positive for SArS-Cov2 by the reference RT-qPCR test. Among these, 262 were positive with the AQ+ InTec test, with a sensitivity of 97.4% (95% CI [95.5-99.3]). The sensitivity increased to 100% (95%CI [100-100]) on samples with a CT level below 25.

In both nasal and nasopharyngeal samples, the sensitivity of the AQ+ InTec test was lowered when the CT level was high. Nevertheless, the sensitivity was always higher with nasopharyngeal samples compared with nasal samples (Table 3).

#### AQ+ InTec test sensitivity in symptomatic and asymptomatic patients

On nasal samples of the 308 symptomatic positive patients, the sensitivity of the AQ+ InTec test was as high as 95.8% and 89.2% on the 83 asymptomatic patients. On nasopharyngeal samples of the 203 symptomatic positive patients, the sensitivity of the test was as high as 97.5% and 97% on the 66 asymptomatic patients.

#### AQ+ InTec test specificity

To evaluate the diagnostic specificity of the AQ+ InTec test, an analysis was conducted on 453 subjects with a negative RT-qPCR reference test who had a nasal sample and 315 subjects with a negative RT-qPCR reference test who had a nasopharyngeal sample.

Two subjects out of 453 (nasal sample) had a positive result with the AQ+ InTec test with a negative RT-qPCR result, giving a specificity of 99.6%.

All patients with a negative RT-qPCR who had a nasopharyngeal sample were also negative with the AQ+ InTec test, giving a specificity of 100% (Table 5).

**Table 4:**
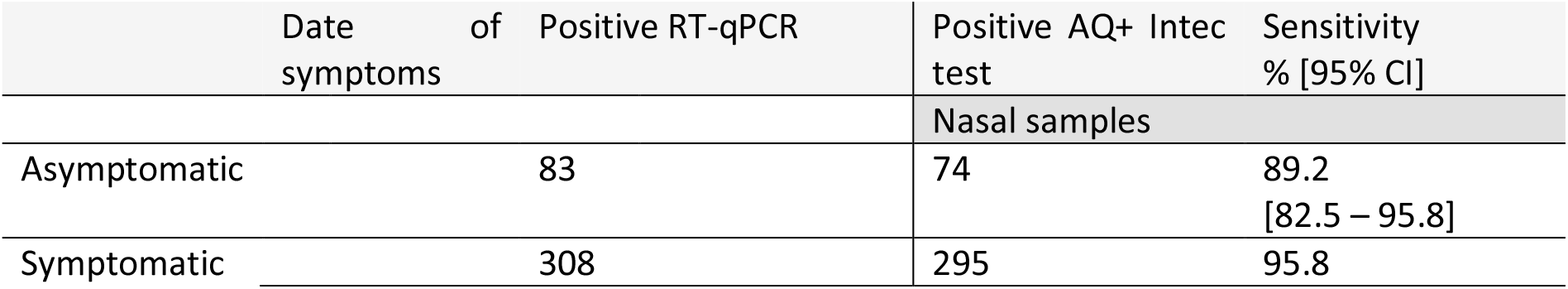
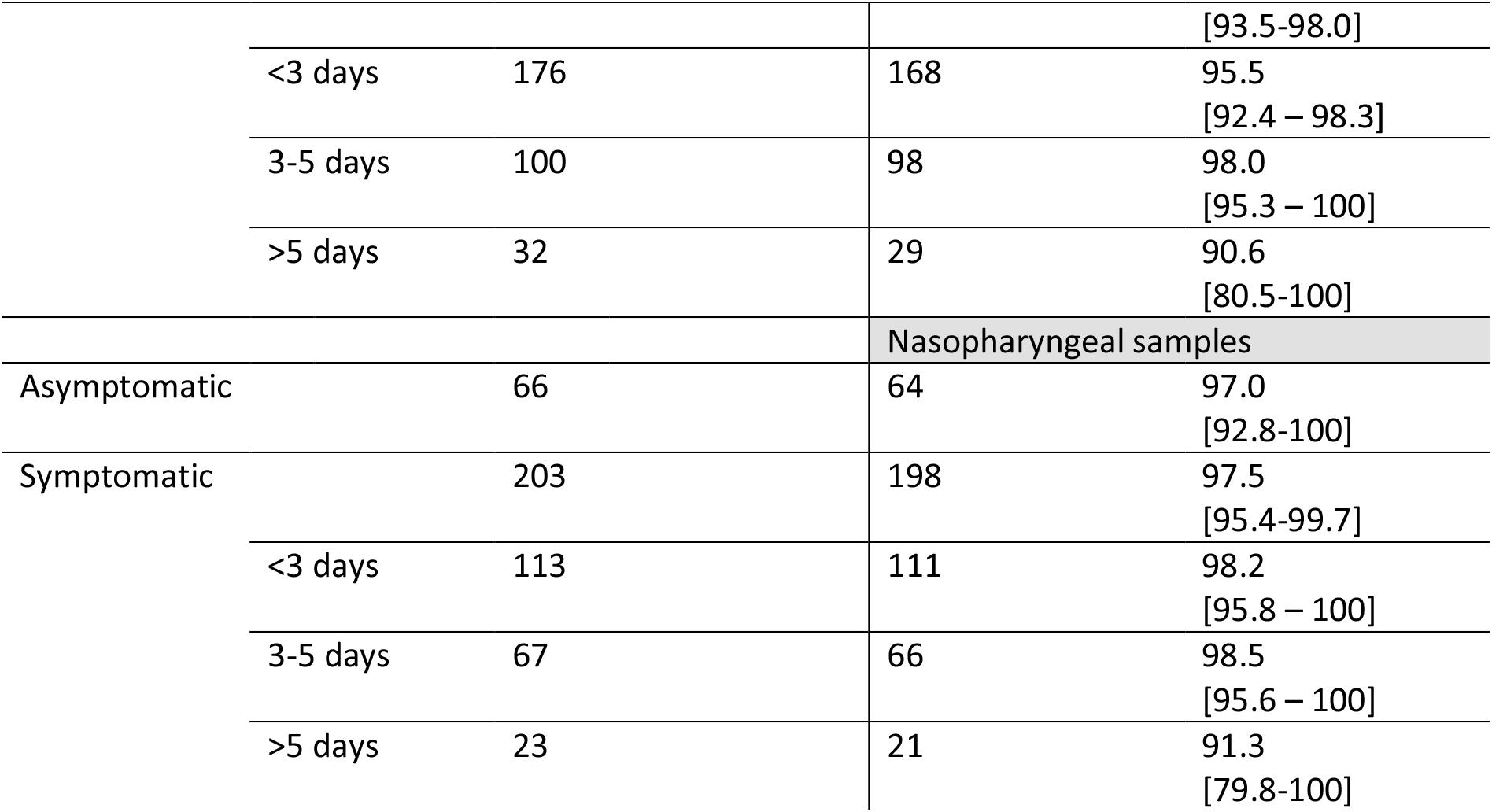
Sensitivity of the AQ+ InTec test on nasal and nasopharyngeal samples in symptomatic and asymptomatic patients

**Table 5:** Specificity of the AQ+ InTec test on nasal and nasopharyngeal samples

| Negative RT-qPCR | Negative AQ+ InTec test | Specificity % [95% CI] |
| --- | --- | --- |
| **Nasal samples** |  |  |
| 453 | 451 | 99.6<br>[99.0 – 100] |
| **Nasopharyngeal samples** |  |  |
| 315 | 315 | 100<br>[100 – 100] |

## Discussion

Retrospective studies on SARS-CoV-2 RADT evaluation allow comparison of different tests on the same sample, as multiple sampling from the same patient raises ethical issues. Indeed, as it is compulsory to use the specific test buffer, it would necessitate a number of swabs equal to the number of RADTs to evaluate for each patient. In this retrospective study, four independent swabs were immersed in a previously collected nasopharyngeal samples, then placed in each of the specific buffers before performing the RADTs allowing statistical comparison, which is not possible in independent studies. However, retrospective studies reduce the real sensitivity because the swab is immersed in 3mL of VTM which dilutes the collected ^19^. The present retrospective study revealed an overall better sensitivity of the AQ+ InTec test compared to the other commercial tests (Orient Gene, Roche, Abott) included in the analysis. Moreover, the LOD analysis revealed a lower LOD for the AQ+ InTec test, followed by Orient Gene, which is consistent with a better sensitivity^20,21^. However, considering the points raised above, these results need to be considered carefully. Of note, in this experimental setup, none of the RADTs, including the AQ+ InTec tests reached a satisfying sensitivity regarding of the acceptance criteria defined in the MDCG 2021-21 (sensitivity >80%)^12^. This lack in performance is easily explained by the experimental setup itself, however, it sheds light on the difference of detection limits that exists between the commercially available SARS-CoV-2 RADT as previously reported^21^. Another concern has emerged regarding the potentially different limits of detection for different SARS-CoV-2 variants between the RADTs^20^. Our study supports that the performance of commercially available RADTs for SARS-CoV-2 detection varies with the variant type, but this should also be considered in perspective with the experimental setup.

In the prospective study aiming at evaluating the sensitivity and specificity test in a point of care setting, the AQ+ InTec test exhibited high sensitivity on nasal samples (94.4%) and nasopharyngeal samples (97.4%) and high specificity (99.6 to 100%) which fulfil the acceptance criteria defined in the MDCG 2021-21 “Guidance on performance evaluation of SARS-CoV-2 in vitro diagnostic medical devices”^12^:

- Diagnostic sensitivity: >80% (rapid tests) relative to the SARS-CoV-2 RT-qPCR test;
- Diagnostic specificity: >98% (rapid tests).

Another point of interest was to compare the results obtained on symptomatic and asymptomatic patients to determine whether the AQ+ InTec test was as sensitive when patients were asymptomatic. Indeed, it has been shown in a study comparing the results of 78 studies on SARS-Cov2 antigenic tests, that the sensitivity of this method is reduced by 13.8 percentage points on average for asymptomatic patients^13–16^.

It is important to note that the sensitivity of the AQ+ InTec test is particularly high on asymptomatic patients compared to other RADTs available on the market. Indeed, an analysis of 12 studies, reported an average sensitivity of 58.1% (95% CI 40.2% to 74.1%)^13^, placing the AQ+ InTec test above in terms of sensitivity on both nasal and nasopharyngeal samples in asymptomatic patients (respectively 89.2% and 97.0%) and making it a valuable tool for testing in an epidemic context with a high proportion of asymptomatic patients that are potential carriers for the SARS-CoV-2 virus ^13–16^.

## Data Availability

All data produced in the present study are available upon reasonable request to the authors

## Supplementary material

**Table S1.**
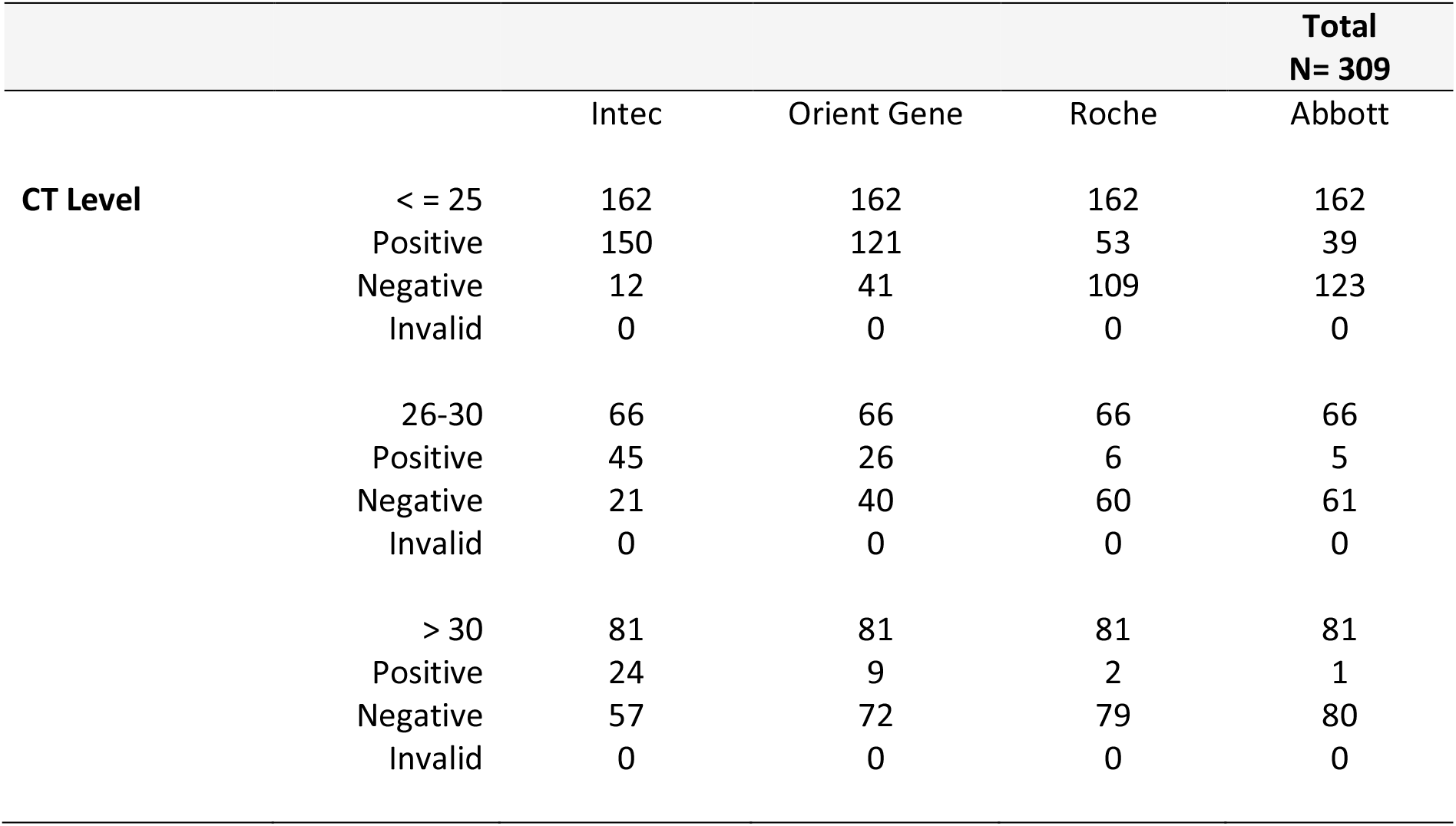
Number of positive RADT according to CT levels obtained with RT-qPCR SARS-CoV-2 detection

|  |  |  |  |  | **Total**<br>**N= 309** |
| --- | --- | --- | --- | --- | --- |
|  |  | Intec | Orient Gene | Roche | Abbott |
| **CT Level** | < = 25 | 162 | 162 | 162 | 162 |
|  | Positive | 150 | 121 | 53 | 39 |
|  | Negative | 12 | 41 | 109 | 123 |
|  | Invalid | 0 | 0 | 0 | 0 |
|  | 26-30 | 66 | 66 | 66 | 66 |
|  | Positive | 45 | 26 | 6 | 5 |
|  | Negative | 21 | 40 | 60 | 61 |
|  | Invalid | 0 | 0 | 0 | 0 |
|  | > 30 | 81 | 81 | 81 | 81 |
|  | Positive | 24 | 9 | 2 | 1 |
|  | Negative | 57 | 72 | 79 | 80 |
|  | Invalid | 0 | 0 | 0 | 0 |

## Aknowledgments

The authors thank Célia Vaslin (Clinact) for providing medical writing and editorial support, in accordance with Good Publication Practice (GPP3) guidelines.

## Author contributions

Experimental strategy design: T. Prazuck, R. Serreau; Experiments: A. Theillay, S. Pallay, D. Pires-Roteia, F. Prazuck, F. Lesne; Manuscript writing: T. Prazuck; Manuscript editing: F. Lesne, T. Prazuck

## Data Availability Statement

The data that support the findings of this study are available from the corresponding author upon reasonable request.

